# The impact of real-time whole genome sequencing in controlling healthcare-associated SARS-CoV-2 outbreaks

**DOI:** 10.1101/2021.04.15.21253894

**Authors:** Rodric V. Francis, Harriet Billam, Mitch Clarke, Carl Yates, Theocharis Tsoleridis, Louise Berry, Nikunj Mahida, William L. Irving, Christopher Moore, Nadine Holmes, Jonathan Ball, C. Patrick McClure, Matthew Loose, on behalf of The COG–UK consortium

**Author notes:** Corresponding author: Professor Matthew Loose, School of Life Sciences, University of Nottingham. https://www.cogconsortium.uk Full list of consortium names and affiliations of the COG-UK members are in the appendix. Joint first authors (Rodric V. Francis, Harriet Billam).

## Abstract

**Background:** Nosocomial infections have posed a significant problem during the COVID-19 pandemic, affecting bed capacity and patient flow in hospitals. Effective infection control measures and identifying areas of highest risk is required to reduce the risk of spread to patients who are admitted with other illnesses. This is the first pandemic where whole genome sequencing (WGS) has been readily available. We demonstrate how WGS can be deployed to help identify and control outbreaks.

**Aims & Methods:** Swabs performed on patients to detect SARS-CoV-2 underwent RT-PCR on one of multiple different platforms available at Nottingham University Hospitals NHS Trust. Positive samples underwent WGS on the GridION platform using the ARTIC amplicon sequencing protocol at the University of Nottingham.

**Results:** Phylogenetic analysis from WGS and epidemiological data was used to identify an initial transmission that occurred in the admissions ward. It also showed high prevalence of asymptomatic staff infection with genetically identical viral sequences which may have contributed to the propagation of the outbreak. Actions were taken to help reduce the risk of nosocomial transmission by the introduction of rapid point of care testing in the admissions ward and introduction of portable HEPA14 filters. WGS was also used in two instances to exclude an outbreak by discerning that the phylotypes were not identical, saving time and resources.

**Conclusions:** In conjunction with accurate epidemiological data, timely WGS can identify high risk areas of nosocomial transmission, which would benefit from implementation of appropriate control measures. Conversely, WGS can disprove nosocomial transmission, validating existing control measures and maintaining clinical service, even where epidemiological data is suggestive of an outbreak.

## Introduction

Severe acute respiratory syndrome coronavirus-2 (SARS-CoV-2) has become a global concern since being first reported in Wuhan, China in December 2019^1^. As of the 28^th^ December 2020, the UK has reported 3,743,734 cases with 103,126 deaths^2^ and 380,839 hospital admissions. The clinical spectrum of COVID-19 infection is wide, ranging from asymptomatic infection to severe viral pneumonia leading to death. SARS-CoV-2 is highly transmissible by droplet and indirect contact^3^. Viral load in asymptomatic patients may be similar to those who are symptomatic^4^. The sensitivity of COVID-19 testing ranges from 71-98% ^5^. This combination of factors poses a significant challenge in infection control.

Nosocomial transmission has been widely reported ^6,7,8^. An early report from Wuhan showed that of 138 hospitalised COVID-19 cases, 12% were identified as being admitted for other reasons and presumed to have acquired the infection in hospital^9^. Transmission between hospital staff has also been demonstrated ^10^. Retrospective analysis in a London teaching hospital showed nosocomial COVID-19 had a case fatality rate of 36%^11^.

At Nottingham University Hospitals (NUH) Trust the Infection Prevention and Control team (IPCT) along with Clinical Microbiology colleagues have hitherto solely relied on epidemiological data to track outbreaks of viral pathogens. Previously whole genome sequencing (WGS) has been used retrospectively to prove/disprove results of outbreak investigations. Coronavirus Disease 2019 (COVID-19) Genomics UK Consortium (COG-UK) was set up in March 2020 to drive WGS nationally^12^, and over 300,000 viral genomes have been sequenced (as of 1^st^ March 2021). This is the first pandemic where WGS technology has been widely accessible in a clinically relevant timeframe, allowing exploration of its utility in a range of clinical scenarios.

We present a series of epidemiologically linked hospital clusters where WGS of SARS-CoV-2 isolates has directly affected real time outbreak management. Sequencing data has been used to both declare outbreaks on inpatient wards as well as stand down and de-escalate possible outbreaks.

## Methods

All patients were tested for SARS-CoV-2 infection on admission to hospital, irrespective of symptomatology. All samples underwent RT-PCR through one of four different molecular platforms (Supplementary information 1).

At the time of analysis, all inpatients with negative admission tests were re-tested every 7 days. Patients who tested negative on admission, but fulfilled clinical criteria for COVID-19 infection were managed similarly to positive patients.

Positive patients were moved to a COVID-19 ward where they were isolated in a side room or cohorted with other positive patients in a bay. Patients negative for COVID-19 were moved to a non-COVID-19 ward. If a bed was not available immediately, patients were isolated or cohorted appropriately on the admissions ward until a bed became available. Type IIR surgical mask, plastic apron and gloves were used as standard personal protective equipment (PPE) on all wards when caring for patients, with enhanced PPE when aerosol generating procedures were carried out. Staff were advised to wear Type IIR surgical masks at all times when inside the hospital which should only have been removed when consuming food or drink.

Symptomatic staff testing was also undertaken. During outbreaks, asymptomatic screening of all healthcare workers who worked in the particular area of concern was also undertaken. Until November 2020, there was no routine asymptomatic healthcare worker testing. The average turnaround time to SARS-CoV-2 results was 11.5 hours from when the swab was received in the laboratory. Results were alerted to the clinical team electronically as soon they were available.

Samples with a positive RT-PCR result and suggestive epidemiological linkage underwent WGS. For routine surveillance of viral sequences not flagged by epidemiological linkage, samples testing positive on the Altona platform with a cycle threshold value of <30 were selected for sequencing. WGS was performed using the ARTIC amplicon sequencing protocol ^13^(Supplementary information 2).

## Results

### Cluster 1

Patients with a diagnosis of a respiratory condition were admitted from both the Emergency Department (ED), as well as directly from the community to a triage respiratory admissions ward, Ward X. A diagnosis of COVID-19 infection was based on a laboratory result, or more rarely on clinical criteria despite negative laboratory testing. Based on the clinical assessment and laboratory results, patients were subsequently transferred to a COVID-19 or a non-COVID-19 ward as appropriate.

Cluster 1 took place on Ward Y; an adult male non-COVID-19 Respiratory ward, consisting of five bays with six beds in each and four side rooms. A timeline for this outbreak is shown in Figure 1.

**Figure 1.**
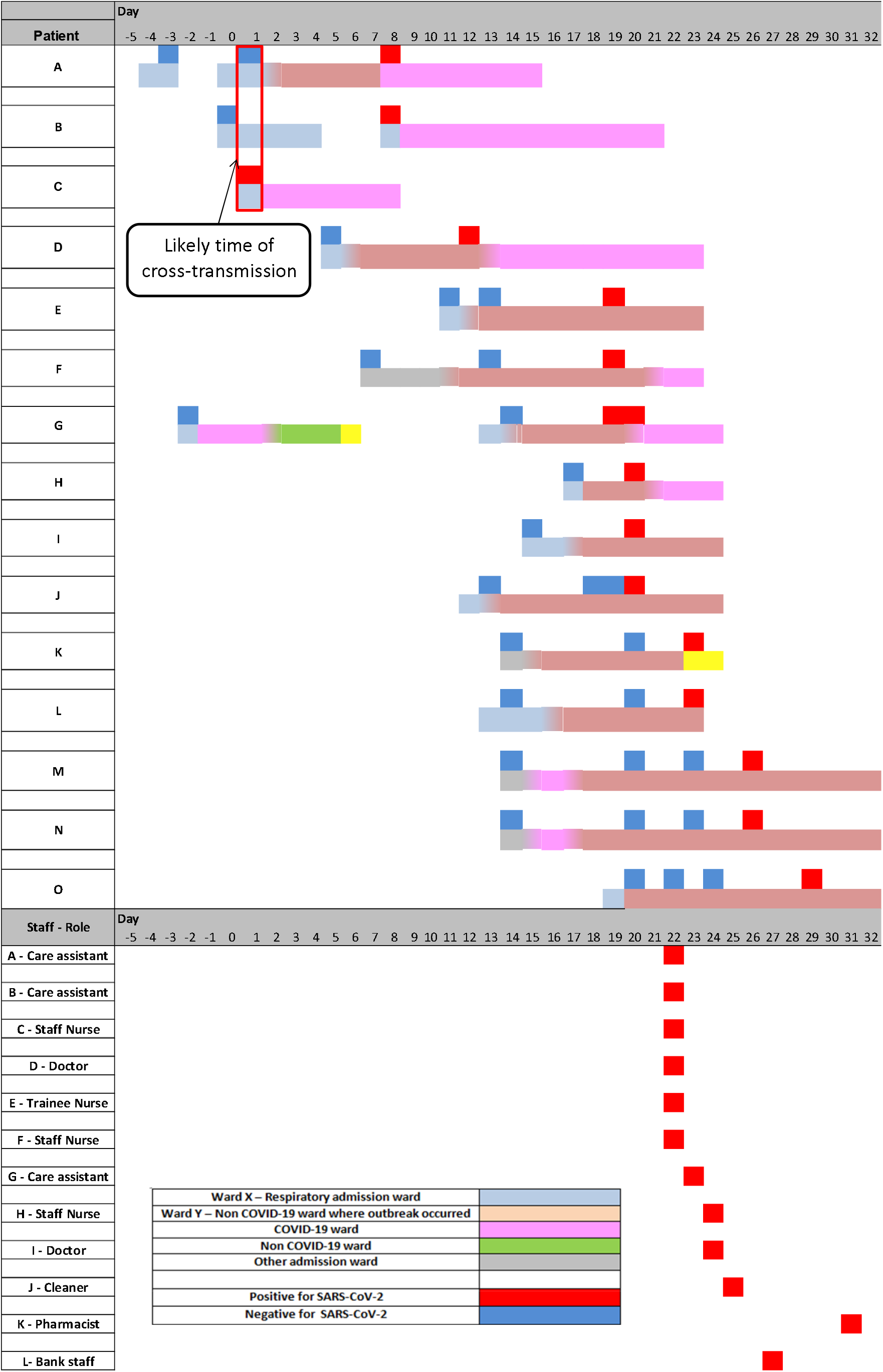
Cluster 1 outbreak timeline demonstrating the cross–transmission event that was likely to have occurred on Ward X which led to multiple cases amongst patients and staff on Ward Y.

Patient A, was admitted to Ward X with breathlessness secondary to a pleural effusion, on Day 0 as defined for the purposes of this cluster. The admission SARS-CoV-2 swab was negative. The patient was transferred to Ward Y 3 days later. On day 8 of admission, the patient developed a fever and worsening hypoxia and a repeat swab detected SARS-CoV-2. The patient was then isolated in a side room before being transferred to a COVID-19 ward.

Patient B was also admitted to Ward X on Day 0 and had a negative result. He was discharged from hospital on Day 4 but subsequently readmitted 2 days later to Ward X, when his admission test detected SARS-CoV-2. WGS results identified Patient A and Patient B’s SARS-CoV-2 genomes as identical.

Patient C was admitted to Ward X on Day 1 with a SARS-CoV-2 induced exacerbation of chronic obstructive pulmonary disease (COPD). Subsequent WGS surveillance identified Patient C to be infected with a genetically identical SARS-CoV-2 virus to Patients A and B (Figure 2). Detailed analysis of the outbreak timeline (Figure 1) in conjunction with the WGS results identified a probable transmission point. All three patients had been located in a bay on Ward X for just 6 hours. This suggested Patient C as the likely index case. This observation is further supported by the later identification of 5 community samples at the base of this branch (Figures 3a and 3b). These community patients were epidemiologically linked to Patient C (Supplementary information 5).

**Figure 2.**
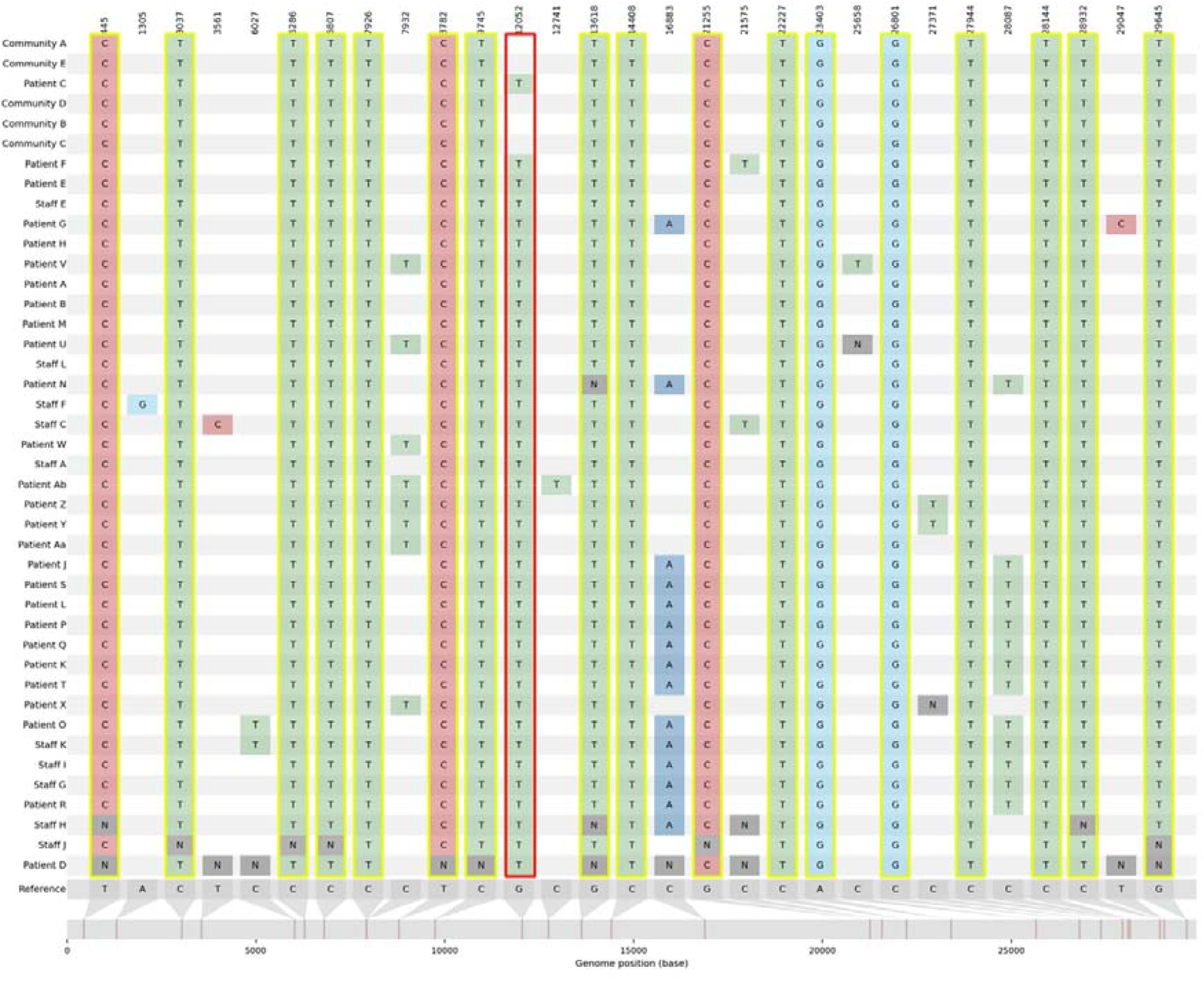
A Snip-it plot from CIVET for samples present in Cluster 1 showing SNPs identified in each sequence. SNPs highlighted in yellow are shared between all samples including those from the community samples. The G12052T SNP is shared between all samples from within the hospital environment.

**Figure 3a.**
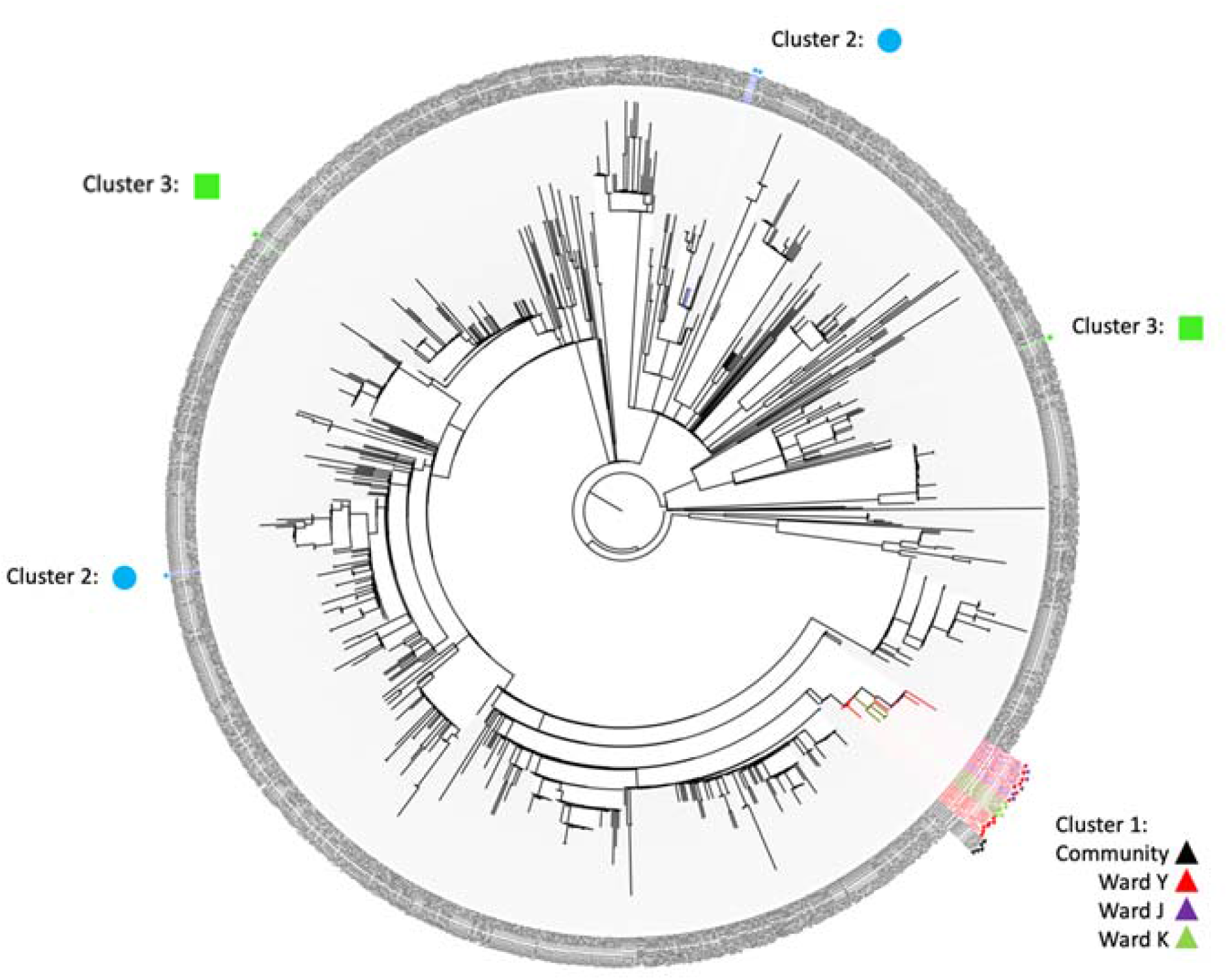
Phylogenetic relationships of SARS-CoV-2 sequences based on their entire genome (29,574nt). The tree represents maximum likelihood phylogenetic analysis of all Nottingham sequences collected between 01/09/2020 and 30/10/2020. Nottingham sequences from hospital Cluster 1 (triangle), Cluster 2 (circle) and Cluster 3 (square) were compared alongside other sequences collected from Nottingham. Hospital isolates that did not meet the >95% coverage criteria were excluded from the analysis. Reference sequences are indicated by their COG accession numbers. Branch lengths are drawn to a scale of nucleotide substitutions per site. Numbers above individual branches indicate SH-aLRT bootstrap support.

**Figure 3b.**
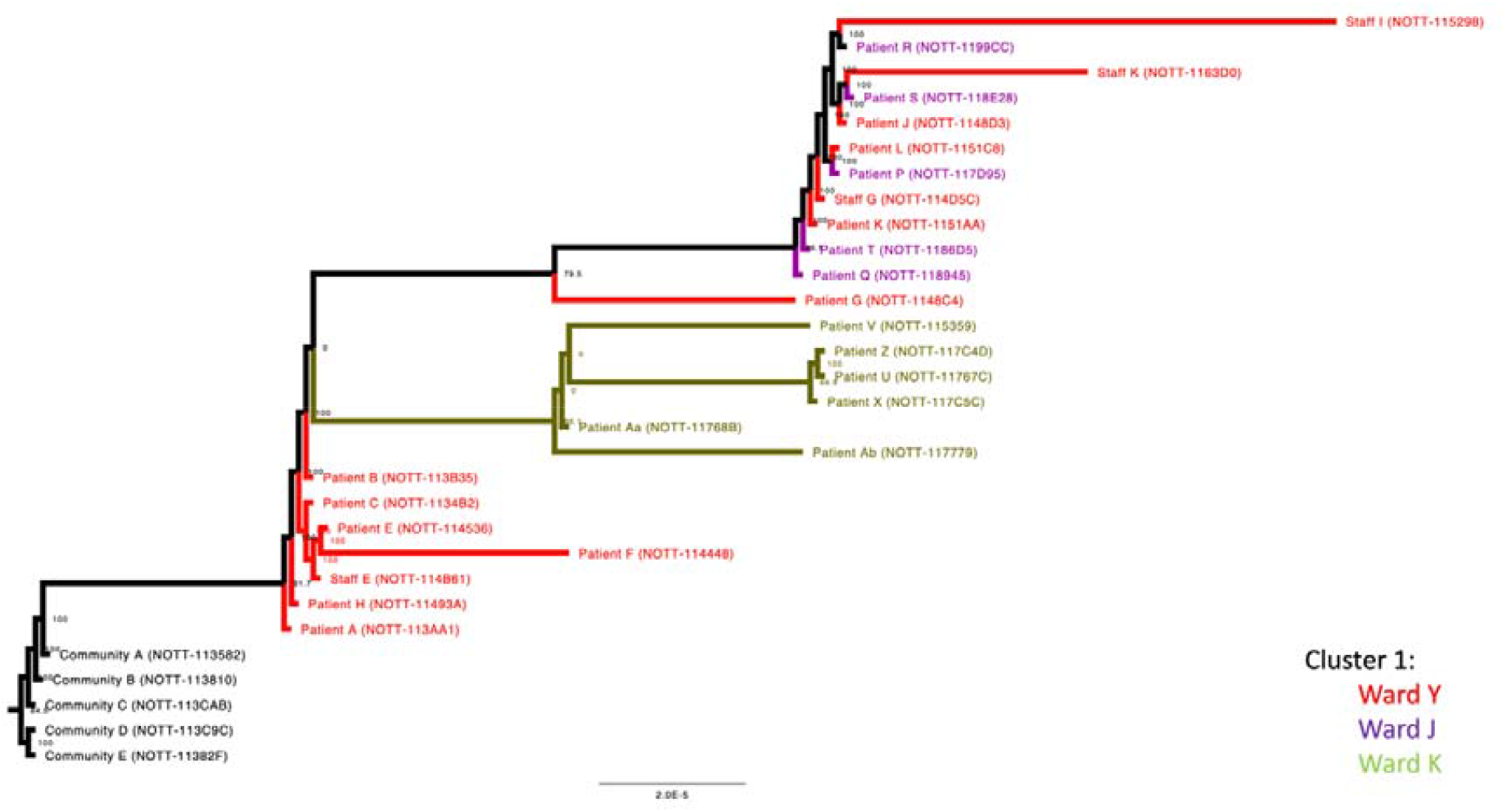
Nottingham sequences from hospital Ward Y (marked in red) were compared alongside sequences from Ward J (marked in purple) and Ward K (marked in green) of the same hospital as well as community sequences (marked in black).

Patient D was admitted to Ward X on Day 1 with pulmonary oedema and an exacerbation of COPD. He tested negative for SARS-CoV-2 and was moved to Ward Y into an open bay. On Day 12 his weekly screening swab flagged positive for SARS-CoV-2. He was promptly moved to a COVID-19 ward. Although only a partial genome sequence was obtained, 14/18 SNPs identified in the viral genome of Patient D matched those of Patients A, B and C with no unique SNPs in this sample.

An outbreak was declared as Patient D had not been a contact of Patient A, B or C. Ward Y was closed for new admissions and asymptomatic screening of patients and staff was undertaken. WGS identified that viruses from Patients E, H and M as well as two staff members (Staff A and E) shared identical SNPs with the original cluster (Supplementary information 5). In total, 8 staff on Ward Y were observed to carry this lineage (3 or fewer differences in SNPs) and were predominantly asymptomatic. This may explain the further spread within Ward Y with no clear contact between the patients.

Five additional patients on Ward Y (Patients J, K, L, N and O) tested positive during Days 20-29. WGS revealed these patients shared all 18 SNPs with the original cluster, but had an additional 2 SNPs (C16883A, C28087T). These SNPs were also found in three healthcare workers from Ward Y (Staff G, H and I). WGS further identified 5 patients on Ward J and 6 patients on Ward K with near identical strains (Supplementary information 5).

Phylogenetic analysis showed that all Cluster 1 isolates formed a distinct clade (Figure 3a). The clade also contained the sub-cluster of 5 community acquired isolates (Community A-E) which were antecedent to all the hospital acquired ones, suggesting this as the route of transmission from community to hospital (Figure 3b). The second sub-cluster comprised 7 Ward Y sequences (Patient A, B, C, E, F, H and Staff E). There were 2 descendant sub-clusters; the first was comprised entirely of 6 Ward K isolates (Patient U, V, X, Z, Aa and Ab), indicating transmission from Ward Y to Ward K. The second one included a mixture of isolates from patients and staff of Ward Y (7 in total) as well as patients of Ward J (5 in total), suggesting multiple transmission events between Wards Y and J.

Overall, the epidemiological data combined with WGS and phylogenetic analysis suggest that nosocomial transmission from a single patient led to clusters of patients and staff positive for SARS-Cov-2 on three wards in the hospital. The primary ward, Ward Y, had 14 patients and 11 staff members positive for SARS-CoV-2. Ward J had 5 patients and Ward K had 6 patients infected as a consequence of this outbreak. The defining SNP for this outbreak, G12052T has been observed in 66 samples in the UK as of 15^th^ December 2020, with 34 of them within this specific outbreak. Given the known context of the community cases surrounding the index patient, this SNP is likely marking this specific outbreak.

The lineage for this strain, B.1.177 has become increasingly prevalent in the UK since its identification^14^ and the specific UK lineage (UK1219) was at least 10-fold more prevalent than any other strain in the East Midlands area as of December 2020, representing approximately 70% of samples identified to date in the area (Supplementary information 4). More recently, B.1.1.7 has displaced this lineage in the East Midlands area.

### Cluster 2

Cluster 2 took place in a renal dialysis unit. Patients in this area were tested for SARS-CoV-2 if they were symptomatic or had been in recent contact with a confirmed positive patient. Patients were allocated an area in the dialysis unit based on their COVID-19 status.

Two cases of SARS-CoV-2 infection were identified in haemodialysis patients attending the same unit. The first, asymptomatic, patient (Patient A2) was screened and tested positive for SARS-CoV-2. Two days later, the second case (Patient B2) attended and tested positive, requiring admission to hospital with hypoxia. An outbreak was declared and screening was performed on 26 patient contacts, all asymptomatic. A third case (Patient C2) was identified from screening which suggested an evolving outbreak. This would have impacted functioning of the dialysis unit, leading to a major operational issue for the Trust. WGS data indicated that the isolates from the first 2 cases were identical but the third case was different (Figure 4). The UK lineage of the shared cases was UK5, whilst the lineage of the individual case was UK1219. This lineage, although similar to that of Cluster 1, is clearly distinct (Figure 3a). The IPCT refocused their investigation on the two identical cases. Patient pathways outside the dialysis unit were reviewed. The only identified connection between them was that both patients travelled together regularly sharing the same ambulance transport to the dialysis unit. A third patient who was also in the ambulance car was negative on both initial screening and on re-testing 7 days later. The driver of the ambulance car was not tested and remained asymptomatic.

**Figure 4.**
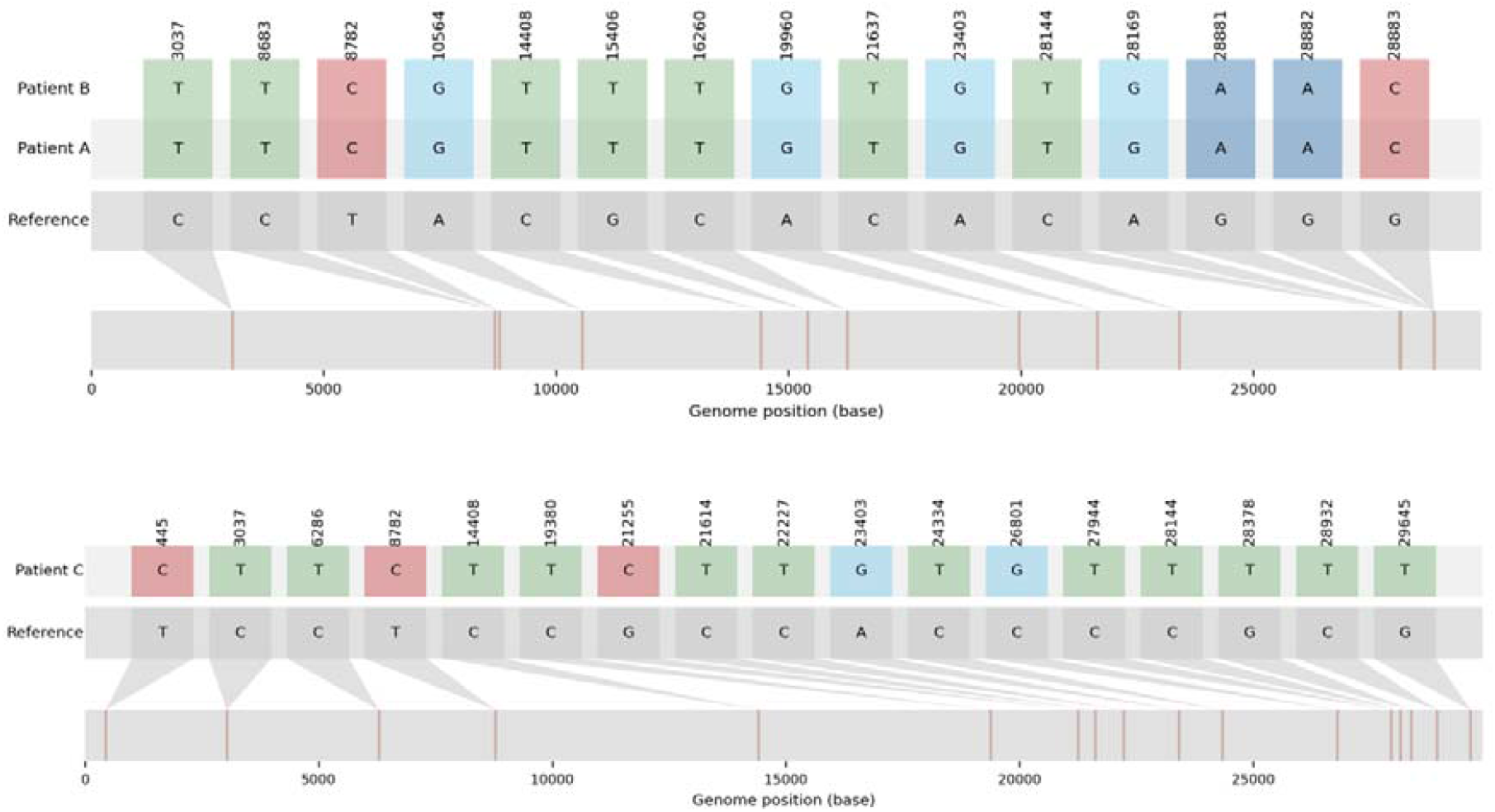
SNP plots showing variants present within the viral genomes derived from the 3 patients in Cluster 2.

A review of the processes around transporting patients to the dialysis unit was undertaken. There was additional focus on appropriate mask usage in the car (type IIR surgical face mask). All patients when interviewed stated that the appropriate mask had been worn at all times in the car. Additional patient education of all patients travelling to the dialysis unit was undertaken in response to this event.

### Cluster 3

Cluster 3 took place on a paediatric general surgical ward, Ward D, which has 11 beds and 4 side rooms. Patients arrive here from a paediatric admissions ward following initial review.

The first case identified (Patient A3) was an infant who had a 58 day inpatient stay following surgical intervention for a prolapsed stoma. During this admission the patient had a negative swab result, and was discharged 8 days later. Nine days following discharge he was readmitted with a positive admission screen for SARS-CoV-2. The previous day a symptomatic staff member (Staff A3) who regularly worked on Ward D tested positive. The possibility of nosocomial transmission was considered, either patient to healthcare worker, healthcare worker to patient, or a third point source as yet undetected, therefore all patients on the ward were screened. This did not identify any further patient cases. No other healthcare workers reported any symptoms. There remained ongoing clinical concerns of a developing outbreak. Wider asymptomatic healthcare worker screening was being considered. Rapid WGS was undertaken, which showed that the patient’s lineage (Patient A3 -UK1219), was very different to the healthcare worker’s strain (Staff A3 – UK352) (Figures 3a and 5). Hence an outbreak was excluded and the ward returned to its normal processes.

**Figure 5.**
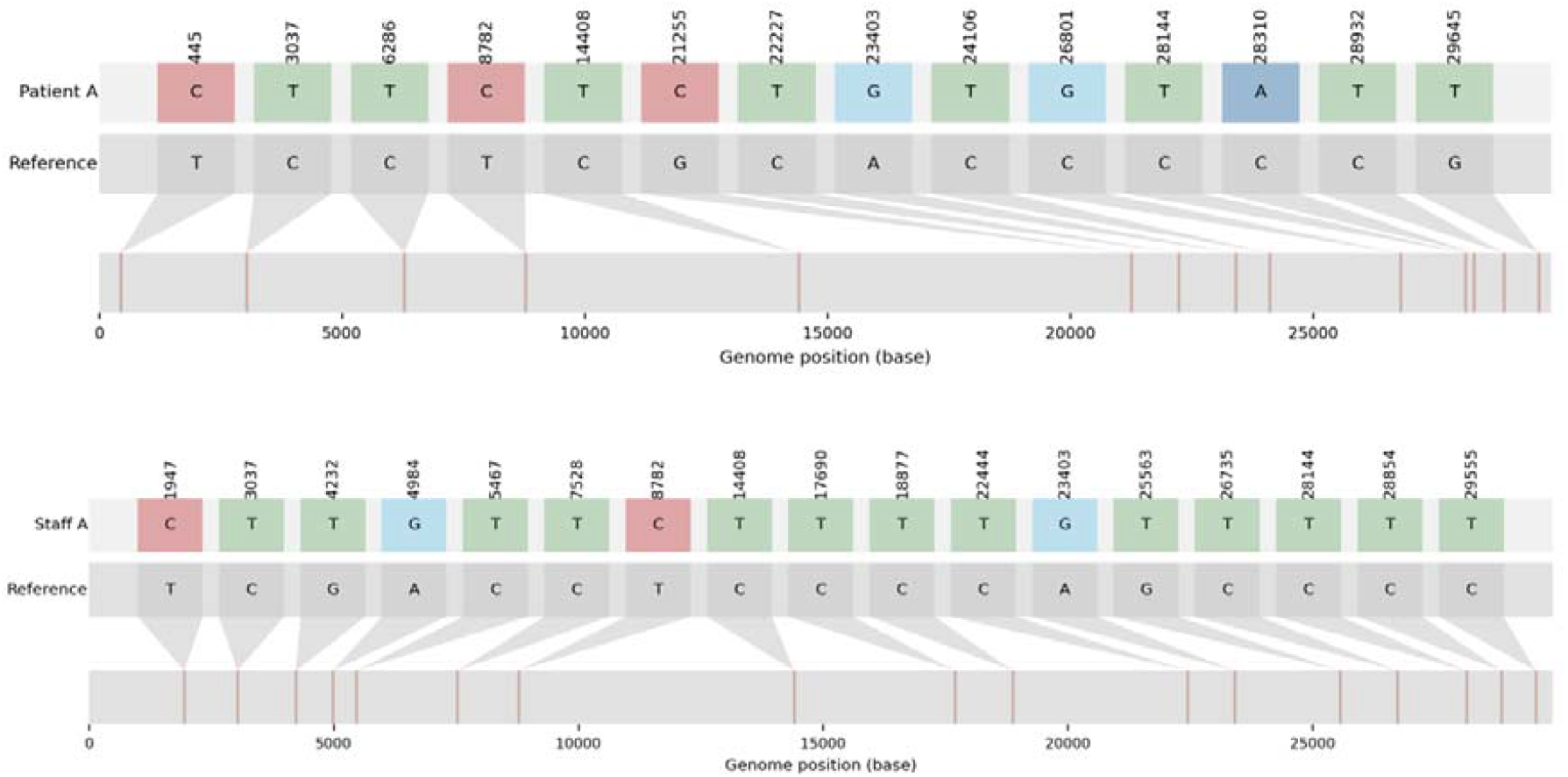
SNP plots for Patient and Staff member in Cluster 3.

## Discussion

The ability to use WGS data at scale to inform hospital outbreaks in close to real time has only been possible in the last decade. Most recently, it has become possible to routinely sequence samples within a 48-hour turnaround and feedback this information to clinical teams whilst an outbreak may be in progress. This has been facilitated by the co-ordinated work of the COG-UK consortium enabling distributed sequencing throughout the UK ^12^. Here we present 3 clusters of SARS-CoV-2 infections which used this rapid feedback mechanism to directly inform active outbreak investigations, allowing the hospital IPCT to focus limited resources on genetically linked outbreaks, which was of particular value during periods of high SARS-CoV-2 positivity.

We have been able to identify transmission occurring in our admission areas using genomics to support the likely route of transmission and exclude other sources in the hospital or the community.

From the examples presented here, the impact of a hospital outbreak is clearly evident. Cluster 1 resulted in numerous infections affecting multiple wards. The ongoing data stream provided by rapid turnaround WGS supplemented the IPCT epidemiological investigation and allowed the identification of additional patients involved in the outbreak. As evidenced by the SNP profiles and phylogeny, patients in Cluster 1 with no direct contact still shared the same lineage. Given that many of the staff who tested positive were asymptomatic, they would have continued working until screening was performed as part of the outbreak management process.

Although the strain (UK1219/B.1.177) was very abundant in the UK at this time (as of early December 2020, accounting for nearly 75% of all cases in the region), this cluster shared a unique SNP specific to this outbreak. In the course of our investigations, we also identified five samples linked to the index patient that had been diagnosed in the community. These five individuals provide the community context by which this lineage entered the hospital but do not contain the SNP unique to the outbreak. There is no evidence at this time that this unique SNP confers any specific phenotype on the lineage. Coupled with the extensive epidemiological data supporting links between these patients and staff, Cluster 1 is a significant hospital outbreak. The application of WGS at the onset of this outbreak enabled the rapid identification of nosocomial transmission and linkage to other wards.

Use of WGS allowed us to identify that transmission of SARS-CoV-2 was occurring in our respiratory admissions area, Ward X. Recognising this allowed us to alter practice quickly, introducing additional control measures. Following this outbreak a Cepheid GeneXpert ® Xpress has been placed on Ward X as well as in the Emergency Department with support of the Point of Care Testing team to further reduce delays caused by transportation of samples. This helps prompt isolation or cohorting of COVID-19 patients, reducing the risk of nosocomial spread to patients who are admitted due to another illness. Air Sentry® HEPA14 air filters have also been deployed onto the admission wards as well as base wards where there has been increased nosocomial transmission. Cluster 1 also highlighted that patients moving between wards can perpetuate ongoing transmission as occurred with Patient K (Supplementary information 2). This cluster also emphasises the importance of future design planning of admission areas to ensure adequate isolation and effective ventilation is available.

In Cluster 2, the WGS results and the epidemiological link between the first two cases, strongly suggested that there had been one cross-transmission event likely to have occurred outside the dialysis unit. This helped close an outbreak investigation quickly, providing assurance that the infection prevention and processes applied within the unit were working well. To date there have been no other cross-transmissions linked to patient transport following additional patient education and emphasis on mask wearing and hand hygiene during patient ambulance transport. This occurrence is similar to that seen in other health care contexts earlier in the pandemic ^8^.

The same applies to Cluster 3, where the clear-cut difference in the strains and weak epidemiological link within the hospital led to the conclusion that the viruses in both patient and staff member had likely been acquired in the community. Prior to WGS, the monitoring process associated with investigation of a potential outbreak would have required considerable time and resources. Cluster 1 and Cluster 2 developed around the same time. WGS results helped focus our efforts on Cluster 1 where nosocomial transmission occurred.

No further cases of epidemiologically or phylogenetically linked SARS-CoV-2 infection were identified in settings related to Cluster 2 and 3 in the weeks following this investigation, further supporting the interpretation of real-time WGS and clinical decisions actioned.

## Conclusion

In conclusion, real-time WGS in association with strong epidemiological evidence has proven to be highly useful in identifying and intervening in hospital SARS-CoV-2 outbreaks and maintaining continuity of service during periods of high viral prevalence, by identifying an outbreak in one of our respiratory admission areas, and disproving nosocomial transmission in 2 other settings despite classical epidemiological methods suggesting linkage.

There is a strong case to be made for continued routine surveillance of infection in local health care settings. Delivering WGS results within 48 hours enables direct response by the IPCT, although even faster turnaround time would have been beneficial in rapid tracing of Cluster 1. These rapid responses can only currently be achieved where sequencing is embedded as closely as possible within the clinical workflow. Implementation and dissemination of WGS in healthcare settings should therefore continue to be supported and strengthened both nationally and globally.

## Data Availability

n/a

## Acknowledgements

*COG-UK is supported by funding from the Medical Research Council (MRC) part of UK Research & Innovation (UKRI), the National Institute of Health Research (NIHR) and Genome Research Limited, operating as the Wellcome Sanger Institute*

## Supplementary data

### Supplementary information 1

Nasopharyngeal swabs were undertaken using Sigma swabs in Virocult^®^ viral transport medium. Upon receipt samples were placed on one of the laboratory multiple work streams based on priority, location and time of day received. Criteria determining which platform was to be utilised for any individual sample were outlined in a Standard Operating Procedure (SOP) through the organisational structures and depended on which area of the hospital the sample was collected from, the need for expediency of the result, and availability of reagents. The platforms used are described below.

#### Work stream 1

Extraction is performed using three different platforms.

- AltoStar^®^ Automation System AM16 (Altona Diagnostics) assay with purification performed using AltoStar^®^ Purification Kit 1.5
- NucliSens easyMAG^®^ extraction system
- Maxwell^®^ RSC Instrument Purification.

Following extraction on one of the above methods, RT-PCR is performed on Bio-Rad CFX96™ Real-Time PCR assay detects SARS-CoV-2 E and S gene targets.

#### Work stream 2

The *m*2000 RealTi*m*e System was used to perform the Abbott RealTi*m*e SARS-CoV-2 assay. This system comprises a sample preparation unit *m*2000*sp*, and amplification and detection unit (Abbott *m*2000rt). This is a dual target assay detecting RNA-dependent RNA polymerase (*RdRp*) and nucleoprotein (*N*) genes.

#### Work stream 3

Cepheid GeneXpert Xpert^®^ Xpress detecting molecular targets *E* and *N* proteins – used for rapid turnaround including emergency surgery or transplant patients.

#### Work stream 4

Novodiag^®^ COVID-19. detecting molecular targets *orf1ab* and *N* proteins – also used for rapid turnaround

### Supplementary information 2

#### Whole genome sequencing

The resulting genomes from the ARTIC amplicon sequencing protocol were aligned and 5’ and 3’ ends were trimmed. The alignment included sequences collected from Nottingham between 01/09/2020 and 30/10/2020 with >95% coverage downloaded from COG-UK (889 sequences). The processing of the genomes was performed using the Geneious Prime 2019.0.4 software. Lineages were assigned to the genomes using the Pangolin tool^15^. A Maximum-Likelihood tree was generated to assess the evolutionary relationships between outbreak sample genomes and other genomes obtained from Nottingham and UK patients. The tree was generated with IQ-TREE2^16^ using the General time reversible model with empirical base frequencies and FreeRate (GTR+F+R2) model of evolution as suggested by the software’s model finder with 1000 SH-like approximate likelihood ratio test (SH-aLRT)^17^. The trees were annotated with Fig tree v1.4.4.

### Supplementary information 3

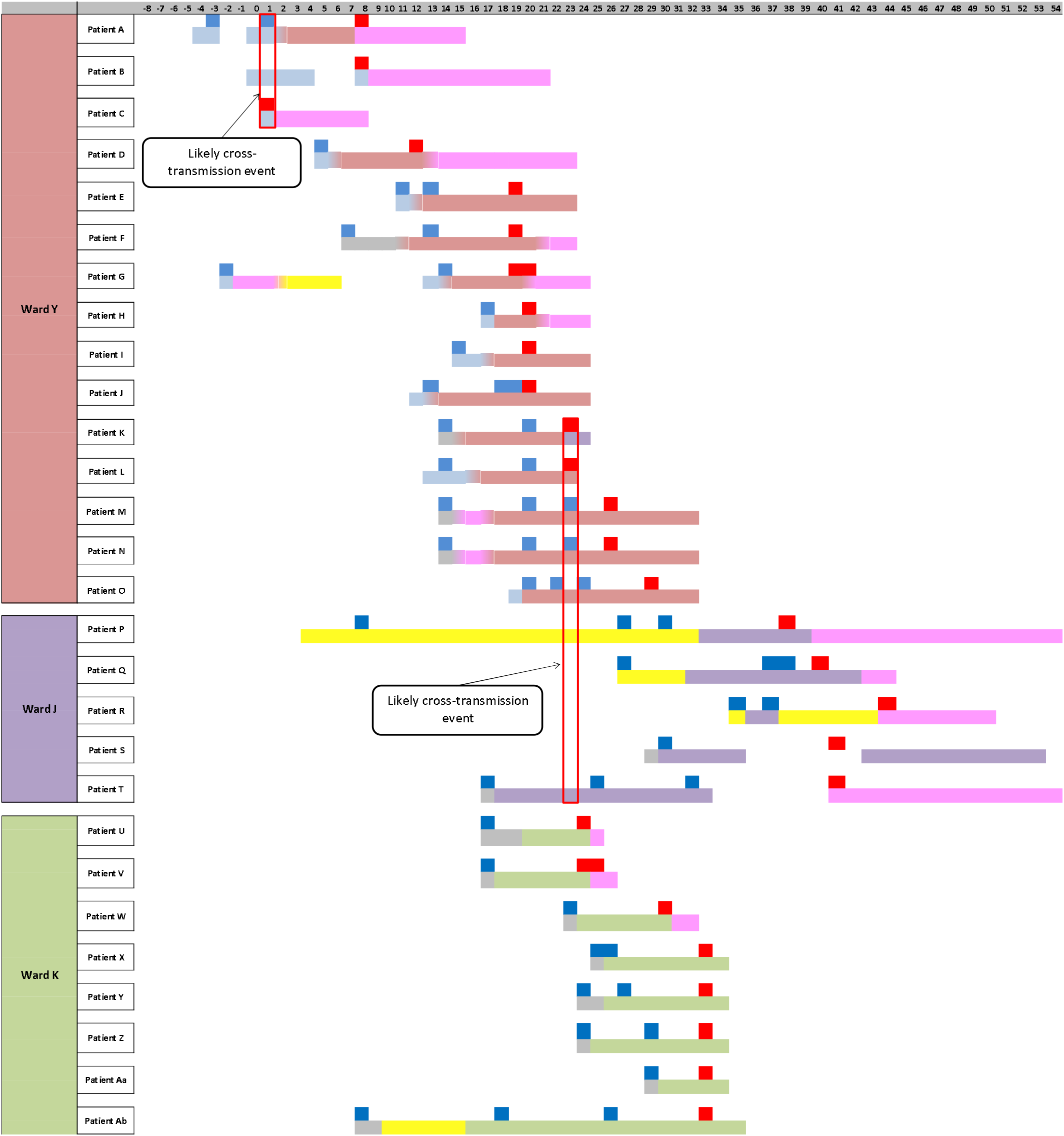

### Supplementary information 4

Table of UK lineages within the NHS England Midlands (North Midland) Region as of 2020-12-15.

| UK Lineage | Count | Global Lineage | Count |
| --- | --- | --- | --- |
| UK1219 | 1668 | B.1.177 | 1660.0 |
| UK1030 | 103 | B.1.1 | 205.0 |
| UK5 | 99 | D.1 | 103.0 |
| UK352 | 78 | B.1.36.1 | 86.0 |
| UK2183 | 56 | B.1.1.37 | 86.0 |
| UK2726 | 52 | B.1.5 | 66.0 |
| UK2397 | 39 | B.1 | 48.0 |
| UK1586 | 38 | B.1.160 | 30.0 |
| UK1614 | 22 | B.1.78 | 22.0 |
| UK1897 | 17 | B.1.1.35 | 13.0 |
| UK3017 | 13 |  |  |
| UK649 | 12 |  |  |
| UK1736 | 11 |  |  |
| UK1805 | 11 |  |  |
| UK2815 | 10 |  |  |
| UK1780 | 9 |  |  |
| UK109 | 8 |  |  |
| UK1506 | 8 |  |  |
| UK1940 | 7 |  |  |
| UK3038 | 7 |  |  |

### Supplementary information 5

#### Additional information relating to WGS of Cluster 1 isolates

Patient C has 1 SNP different to the community samples, namely G12052T. Thus a set of 18 SNPs were identified in common in all subsequent individuals in Cluster 1 (see Figure 1, 2). This cluster is from pangolin lineage B.1.177 and UK lineage UK1219 with the phylotype UK1219_1.76.2.1.2.2.5.34.1.1.9.1.

WGS identified that Patients E, H and M as well as two staff members (Staff A and E) shared identical SNPs with the original cluster. For one member of staff (Staff J) WGS only generated a partial genome, but again the SNPs that could be called (13/18) were identical to the main cluster. Patients F and G were separated by 1 SNP (C21575T) and two SNP’s (C16883A, T29047C) respectively. Although Patient I tested positive, WGS was unsuccessful for this patient.

Amongst the remaining staff on the ward, two staff members harboured viruses which differed by 1 SNP (Staff F and H), two staff by 2 SNPs (Staff C, G and I) and 1 staff by 3 SNPs (Staff K). All these staff members shared the core set of SNPs. Virus from one remaining staff member (Staff D) only shared 12/18 SNPs with this cluster with an additional 3 SNPs with respect to the reference strain giving a total of 9 nucleotides difference. As a consequence, this individual was not considered part of this cluster. Within the wider hospital staff community only one other staff member, Staff L, had a virus of the same lineage as the cluster and shared 18/18 SNPs.

WGS further identified 5 patients on Ward J and 8 patients on Ward K with near identical strains (See Supplementary data 4). These samples held all SNPs in common with samples from Patients P, Q, R, S and T from Ward J. Patient P’s positive result predated the variants found on Ward Y. Ward J is a 20 bedded adult renal medicine ward with 4 side rooms and 4 bays with 4 beds in each bay. Patient K was moved from Ward Y to Ward J on Day 23 (see Figure 1) for 48 hours. During this time Patient T was also present on this ward. This identified a potential cross transmission event on Ward J. (see Supplementary information 2)

Another link was identified by WGS on Ward K, a 28 bedded care of the elderly ward with 4 side-rooms and 4 bays with 6 beds in each bay. Again, 8 patients (Patients U, V, W, X, Y, Z, Aa and Ab) were found to share all 18 SNPs with the original cluster. All 8 patients shared an additional SNP (C7932T). Patients V, Y, Z and Ab each had an additional SNP suggesting further within-ward transmission. No staff members on Ward Y held this specific subset of SNPs.

